# A cross-sectional survey of material deprivation and suicide-related ideation among Vietnamese technical interns in Japan in 2021

**DOI:** 10.1101/2022.12.14.22283455

**Authors:** Tadashi Yamashita, Pham Nguyen Quy, Chika Yamada, Emi Nogami, Erina Seto-Suh, Saori Iwamoto, Kenji Kato

## Abstract

**Background:** Many technical intern trainees in Japan are economically impoverished because of the need to send money back to their home country, debts from the intermediaries that arranged their arrival in Japan, and reduced working hours because of the coronavirus disease 2019 (COVID-19) pandemic. In addition, there is concern that COVID-19 may cause mental instability in response to the life changes experienced by interns. The purpose of this study was to elucidate the experience of material deprivation and the relationship between the state of material deprivation and suicidal ideation among Vietnamese intern trainees in Japan.

**Methods:** A cross-sectional study was conducted from September to October 2021. Of 310 Vietnamese technical intern trainees who responded, we analyzed data from 200 individuals with no missing or abnormal values. The questionnaire obtained information about gender, age, length of residence in Japan, Japanese language proficiency, changes in income related to the COVID-19 pandemic, material deprivation status, and suicidal ideation. The ninth item of the Patient Health Questionnaire-9 was used to examine suicidal ideation. Logistic regression analysis was used to analyze the relationship between material deprivation items and suicidal ideation.

**Results:** Respondents’ mean age was 26.0 ± 5.1 years, and 62.0% (n = 124) were male. Regarding material deprivation items, food was reported in 82 (41.0%) cases, cellphone bills were reported in 49 (24.5%) cases, and medical expenses were reported in 34 (22.0%) cases. Forty-six (23.0%) respondents reported experiencing suicidal ideation, and the prevalence was associated with age (p = 0.031, odds ratio [OR] = 0.889, 95% confidence interval [CI] = 0.799–0.990), deprivation regarding food expenses (p = 0.003, OR = 3.897, 95% CI = 1.597–9.511), and deprivation regarding cellphone usage (p = 0.021, OR = 3.671, 95% CI = 1.217–11.075).

**Conclusions:** Vietnamese technical intern trainees in Japan experienced material deprivation in multiple ways, and exhibited a high prevalence of serious psychological problems. Factors contributing to suicidal ideation included age, experience of deprivation in relation to food expenses, and deprivation in relation to cellphone bills. Inability to pay cellphone bills may have increased isolation among Vietnamese trainees during the COVID-19 pandemic.

## Background

The World Health Organization reported that the coronavirus disease 2019 (COVID-19) pandemic has led to an increase in the prevalence of depression worldwide, with social isolation being a major factor.[1] In addition, migrants are reported to be at an increased risk of mental health problems during the COVID-19 pandemic, because of stress related to their concerns about the health of their family and friends, economic problems, and uncertainty about their future.[2] To support migrants’ health, it is necessary to improve access to necessary health services and to address linguistic and socio-cultural barriers. It is particularly important to provide livelihood and health support in terms of social health disparities for migrants who are at high risk of social isolation because of the impact of the COVID-19 pandemic.

At the end of 2021, the number of foreign residents in Japan was approximately 2.76 million, of which Vietnamese residents accounted for 15.7%. In terms of the population of foreigners in Japan by nationality, Chinese people account for the largest proportion, while Vietnamese people account for the second-largest proportion. Regarding the residence status of Vietnamese residents, the most common status is technical intern, which accounts for approximately 40% of Vietnamese residents in Japan.[3] The number of Vietnamese technical interns in Japan tripled between 2015 and 2019. This situation emerged because of an increase in the number of opportunities for Vietnamese technical trainees to work in Japanese industries and to earn a salary to support their families living in Vietnam.[4] However, in addition to the physical and mental stress of working in an unfamiliar country, Vietnamese technical intern trainees face many obstacles, including severe restrictions on their right to choose their occupation and where they can immigrate. Human rights violations have been reported in Japan, including the disappearance of Vietnamese technical intern trainees, verbal abuse of Vietnamese technical intern trainees by the facilities receiving them, and dismissal of Vietnamese technical intern trainees for pregnancy.[5,6,7] The Ministry of Justice in Japan has taken steps to prevent the recurrence of these problems. Additionally, a previous study reported that Vietnamese female technical intern trainees working in Japan were often unable to talk to anyone about their health problems, which worsened their condition.[8] These findings suggest that some technical intern trainees in Japan face severe social and health conditions.

Migrants are more likely to experience social deprivation,[9] and it has been reported that those who experience material deprivation are more likely to develop depressive symptoms and mental health disorders.[10][11] With the outbreak of the COVID-19 pandemic, technical intern trainees working in Japan suddenly had fewer opportunities to work because of reduced working hours and layoffs. This has had a significant impact on interns’ economic livelihood.[12,13] In addition, there have been reports of cases in which the intermediaries operating between employers and technical intern trainees working in Japan have overcharged for intermediary fees, financially exploiting the interns.[14,15] Thus, Vietnamese trainees in Japan may have experienced material deprivation as a result of the COVID-19 pandemic, potentially causing mental health problems. However, no previous studies have examined the conditions experienced by Vietnamese trainees in Japan during the COVID-19 pandemic regarding their experiences of material deprivation and their mental health. Therefore, the current study aimed to clarify the current situation of Vietnamese technical intern trainees’ experiences of material deprivation and their depressive symptoms and suicidal ideation. Furthermore, we sought to clarify the extent to which Vietnamese technical intern trainees’ experiences of material deprivation affect the prevalence of suicidal ideation.

## Methods

This study used a cross-sectional research design with an online questionnaire. The study was conducted in accord with the Declaration of Helsinki and was approved by the Ethics Committee of Kobe City College of Nursing (approval number: 20124-05). Participants read an explanation of the purpose of the study, which explained that participation was voluntary, that participants would not be disadvantaged by non-participation, and that they would not be personally identified before answering the questionnaire. Subsequently, after the participants agreed to participate, they were enrolled in the study.

### Participants

310 responses were received from Vietnamese technical intern trainers in Japan. In the current study, 200 of the 310 responses were included in the analysis, excluding responses with missing and outlier values.

This web-based survey was conducted using SurveyMonkey (Momentive Inc., San Mateo, CA, USA). All instructions for obtaining consent to participate in the survey, as well as all questions and answers within the survey, were written in Vietnamese. Participants were recruited by posting recruitment notices in groups of Vietnamese people living in Japan via Facebook (Meta Platforms Inc., Menlo Park, California, USA). The questionnaire in SurveyMonkey was set to not allow duplicate responses. Participation requirements were used to select Vietnamese nationals currently residing in Japan, who were 18 years of age or older. Exclusion criteria were used to exclude those who could not read the participation instructions and those who could not answer the questions by themselves. After responding to the survey, we sent an online coupon worth 200 Japanese yen to the registered e-mail address as a reward for participating in this study.

### Measures

The surveyed characteristics of participants were age, length of residence in Japan, how many people they lived with, gender, residence status, educational background, marital status, level of Japanese language proficiency, and current medical history. Social and economic items within the questionnaire were health insurance, availability of health advisors, and methods used for obtaining information relevant to their daily lives. Additionally, the questionnaire collected information about changes in monthly income and employment status during the COVID-19 pandemic. Furthermore, we obtained information about participants’ experiences of material deprivation after entering Japan. The respondents were asked about their current income compared with before the COVID-19 pandemic, using a four-case method (decrease of 40% or more: significant decrease; decrease of 10%–40%: slight decrease; decrease of less than 10%: almost the same; and increase of 10%–40%: slight increase). Material deprivation was defined by the Japanese Cabinet Office as the lack, for economic reasons, of goods and services that are typically available and enjoyed in the country in which one lives.[16] Seven items of material deprivation published by the National Institute of Population and Social Security

Research were used in this study.[17] The material deprivation indicator asked about experiences of not being able to pay or purchase goods while in Japan. Respondents were instructed to answer using four response options: never, rarely, sometimes, or often. The Patient Health Questionnaire-9 (PHQ-9) was used to measure the degree of clinical symptoms of depression. The PHQ-9 measures the degree of clinical depressive symptoms and has four levels (not at all, several days, more than half of days, and nearly every day).[18] The Vietnamese version of the PHQ-9 was used in this study.[19,20] The internal reliability of the PHQ-9 used in this study was confirmed, with a Cronbach’s alpha coefficient of 0.90. To ascertain suicidal ideation, the ninth item of the PHQ-9 was used. When assessing suicidal ideation, one item from the depression scale can be taken and used as a scale to assess suicidal ideation.[21] This method has been used in several previous studies.[22,23,24] The ninth item on the PHQ-9 was as follows: “Thoughts that you would be better off dead or thoughts of hurting yourself in some way?” In this survey, “not at all” was scored as 0, “several days,” “more than half of days,” or “almost every day” were scored as 1. We considered 1 to indicate suicidal ideation.

### Statistical Analysis

In the analysis, descriptive statistics for each variable were first calculated. After checking for multicollinearity between variables, logistic regression analysis using the forced entry method was conducted to examine the relationship between suicidal ideation and material deprivation of Vietnamese technical interns in Japan. Confounding factors for the logistic regression analysis were selected with reference to several previous studies, and these were characteristics of participants and their socioeconomic status. [25][26][27] [28]

In the logistic regression analysis, the level of Japanese language proficiency was transformed into a binary variable (0 for fluent Japanese and 1 for less fluent Japanese). Change in monthly income was also transformed into a binary variable (0 for no change or increase and 1 for decrease). Additionally, material deprivation was converted into binary values (0 for no deprivation and 1 for a few days, more than half a day, and almost every day). However, among the items of material deprivation, because the gas and electricity bills are often paid together in Japan, the variables for the gas and electricity bills were combined and scored as 1 if the respondent was unable to pay either the gas or electricity bill, and 0 otherwise. A total of six items of material deprivation were entered into the logistic regression analysis. All statistical analyses were performed with SPSS version 19 software (IBM Corp., Armonk, NY, USA). Statistical significance was set at P < 0.05 (two-sided tests).

## Results

Participants’ mean age was 26.0 ± 5.1 years, the mean number of years living in Japan was 2.9 ± 4.4 years, and the mean number of people living together was 3.9 ± 2.7 years. 124 (62.0%) participants were male and 76 (38.0%) participants were female. The most common educational background status was high school (112 [56.0%]), followed by technical school (47 [23.5%]). 167 (83.5%) participants were single. Regarding the level of Japanese language proficiency, 10 (5.0%) participants could speak fluent Japanese, 83 (41.5%) could speak enough Japanese to not affect their work or study, 81 (40.5%) could speak enough Japanese to not affect their daily life, and 26 (13.0%) could barely speak Japanese. 41 (20.5%) participants had pre-existing health conditions.

68 respondents (34.0%) had someone they could consult regarding their health. The most common method of obtaining information relevant to daily life (multiple choice) was social networks such as Facebook (164, 82.0%), followed by website searches such as Google (91, 45.5%), Vietnamese friends (85, 42.5%), television, newspapers, and magazines (55, 27.5%), and at work or school (35, 17.5%).

Regarding changes in monthly income during the COVID-19 pandemic, 71 (35.5%) participants reported a significant decrease in income of 40% or more, 91 (45.5%) participants reported a slight decrease of 10%–40%, 35 (17.5%) reported a change of less than 10%, and three (1.5%) participants reported a slight increase of 10%–40%. Employment status during the COVID-19 pandemic was reported as dismissal or unemployment by 15 (7.5%) participants. Change in employment status was reported by nine (4.5%) participants, change in type (content) of work was reported by 19 (9.5%) participants, and reduction in working hours and days was reported by 142 (71.0%) participants.

The items in which material deprivation was observed were as follows. Food shortages were experienced “frequently” by four (2.0%) and “sometimes” by 78 (39.0%) participants. Clothing shortages were reported to occur “frequently” for 14 (7.0%) participants and “sometimes” for 67 (33.5%) participants. Difficulty paying cellphone bills was experienced “frequently” by 10 (5.0%) participants and “sometimes” by 39 (19.5%) participants. Electricity bills were in arrears “frequently” for three (1.5%) participants and “sometimes” for 37 (18.5%) participants. Gas bills were reported to be in arrears “frequently” by four (2.0%) participants and “sometimes” by 35 (17.5%) participants. Rent arrears were reported to be paid “frequently” by nine (4.5%) participants and “sometimes” by 30 (15.0%) participants. Eight respondents (4.0%) “could not pay frequently” and 36 (18.0%) “could not pay sometimes” regarding deprivation of medical expenses.

**Figure 1.**
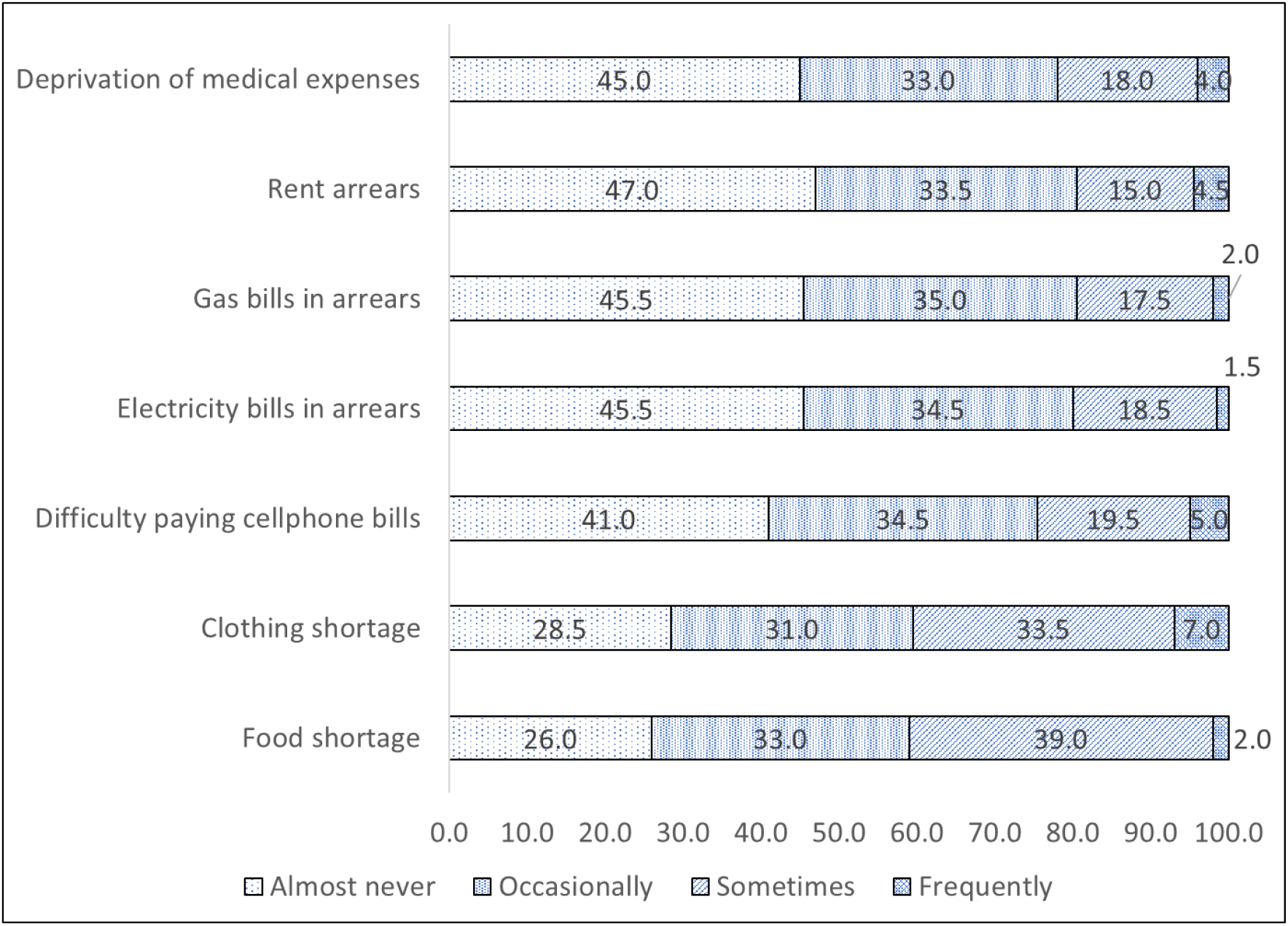
Material deprivation status among Vietnamese technical interns in Japan in 2021 (n = 200)

The PHQ-9 score was 7.9 ± 6.2. Severe (15–19 points) and very severe (20–27 points) depressive symptoms were reported by 17 (8.5%) and 12 (6%) participants, respectively.

The number (percentage) of participants scoring 1 or more points (with suicidal ideation) on the nine items of the PHQ-9 was 46 (23.0%).

The results of the single regression analysis revealed significant differences in age (P = 0.016, odds ratio [OR] = 0.890, 95% confidence interval [CI] = 0.810–0.978), food deprivation (P = 0.001, OR = 3.251, 95% CI = 1.639–6.448), and deprivation regarding cellphone bills (P = 0.003, OR = 2.909, 95% CI = 1.431–5.914). The items that showed significant differences in the multivariable logistic regression analysis were age (P = 0.031, OR = 0.889, 95% CI = 0.799–0.990), food deprivation (P = 0.003, OR = 3.897, 95% CI = 1.597–9.511), and deprivation regarding cellphone bills (P = 0.021, OR = 3.671, 95% CI = 1.217–11.075).

## Discussion

The survey conducted at the end of 2021 revealed that some Vietnamese trainees living in Japan had difficulty paying for food, clothing, cellphones, electricity, gas, and rent. Approximately 20% of the trainees surveyed had high suicidal ideation, indicating that some Vietnamese trainees have serious psychological difficulties. Items associated with suicidal ideation were age, food deprivation, and deprivation of income to pay cellphone bills. Inability to pay cellphone bills may have increased isolation among Vietnamese trainees during the COVID-19 pandemic. As the COVID-19 pandemic continues, a system for early detection and support of Vietnamese trainees with suicidal ideation is required. Additionally, it is important to provide financial support for Vietnamese technical intern trainees and prevent their isolation.

The economic burden on Vietnamese technical intern trainees in Japan to continue their socioeconomic activities in Japan has increased as a result of the COVID-19 pandemic. The Japanese Ministry of Health, Labour and Welfare has been providing assistance to technical intern trainees who have difficulty working because of the deterioration of their employer’s management caused by new coronavirus infections, so that they can change their status of residence and remain in Japan.[29] Meanwhile, the deterioration of the socioeconomic situation of technical intern trainees and the resulting anxiety are becoming increasingly serious problems because of prolonged infection with new types of coronavirus. A report from a hotline consultation for technical intern trainees conducted by the Japan Catholic Committee for Refugees, Migrants and Migrants[30] revealed that the COVID-19 pandemic has halved the amount of available work, drastically reduced wages, and left interns with no one to consult even when they have serious health concerns.[31] Supporting the health of Vietnamese technical intern trainees is a concern, as foreign residents in Japan have been reported to have inadequate access to medical care in Japan.[32]

The effects of new coronavirus infections may also impact the health of young technical interns because of their unfamiliarity with life in a foreign country. An online survey of 589 Vietnamese residents in Japan from January to March 2021 indicated that 37.2% of technical interns are depressed, exhibiting Center for Epidemiologic Studies Depression Scale scores of 16 or higher.[33] Although the timing of the current survey and this previous study differ, the approximate percentages are similar. The results of a survey conducted in October 2021 of 3,262 Japanese technical intern trainee supervisory organizations reported that more than half of the organizations were struggling to respond to consultations regarding mental illness among technical intern trainees.[34] The repeated declarations of a state of emergency in Japan since 2020 under the COVID-19 pandemic may have reinforced anxiety among young Vietnamese technical intern trainees who are engaged in social activities away from their home countries. In a survey conducted using the same scale among Japanese university students in May–June 2020, the rate of suicidal ideation was 6.7%.[23] Thus, the rate of suicidal ideation among Vietnamese technical intern trainees was found to be higher than that of Japanese university students under the COVID-19 pandemic. This suggests that the outbreak of the novel coronavirus may have placed a stronger psychological burden on young Vietnamese technical interns compared with Japanese university students who were accustomed to living in Japan.

In the current study, the material deprivation situation of Vietnamese trainees was also found to contribute to suicidal ideation. A South Korean panel study using data from 2012–2018 shows an association between relative deprivation of income and likelihood of suicide attempts,[35] which is similar to the present results. Furthermore, the items found to be associated with suicidal ideation among Vietnamese technical intern trainees in this study were found to be related not only to the experience of being deprived of food necessary for daily life in Japan, but also to the experience of being deprived of cellphones. Several theories exist regarding the link between suicidal ideation and cellphones. One theory is that prolonged use of cellphones promotes isolation among young people, leading to suicidal ideation.[36,37] Another theory is that cellphones have reduced isolation and improved mental health among young people during the COVID-19 pandemic. Studies supporting the latter theory include a 2022 survey of Japanese university students revealing that smartphone use reduced the risk of loneliness during a pandemic,[38] a study conducted in Belgium during the COVID-19 pandemic that showed that those who felt lonely were more likely to use social media to cope with a lack of social contact,[39] and a study conducted in Australia during the COVID-19 pandemic reported lower levels of adolescent loneliness among those who used technology to connect with society.[40] Thus, in the extraordinary situation of the COVID-19 pandemic, it is possible that young people were not dependent on social media, but rather used it as a tool to combat loneliness. This may also apply to Vietnamese technical interns who have left their home country to live in Japan. Because the majority of trainees in the current study were in their early 20s, it is likely that many of them were in contact with family members and distant friends in their home countries via social media at the time of the outbreak of the COVID-19 pandemic. During the COVID-19 pandemic, cellphones were likely to be an important tool for technical interns to connect with people in their home country and provide mental comfort. This study revealed that deprivation of this important tool had a significant impact on participants’ mental state.

In the future, health centers and public health centers in areas in which Vietnamese technical intern trainees reside should cooperate with technical intern training organizations and employers to establish a system for early detection of Vietnamese technical intern trainees with suicidal tendencies. It will also be necessary to consider remote support using social networking sites and other means to ensure that Vietnamese technical intern trainees are not isolated during the COVID-19 pandemic. Furthermore, it is necessary to provide financial support as well as psychological support to ensure that Vietnamese technical intern trainees living in Japan do not become materially impoverished.

The current study involved several limitations. First, the measures of material deprivation and depression were obtained through subjective responses, which may have involved cognitive bias. Second, recruitment of participants through Facebook may have created a selection bias for individuals with a high level of interest in this study. Additionally, in this study, we were unable to take into account the emotional stress felt by the Vietnamese trainees in Japan as a result of being away from their home country. Finally, because the data in this study were collected after the COVID-19 pandemic, comparisons with data collected before the outbreak of novel coronavirus infection were not possible. However, we believe that this survey has clinical significance in that it obtained insights regarding the mental health of Vietnamese technical intern trainees in Japan using objective indicators.

## Conclusions

The purpose of the current study was to determine the actual conditions of deprivation and mental health of Vietnamese technical intern trainees during the COVID-19 pandemic and to examine factors related to suicidal ideation. The results of this study indicated that Vietnamese technical intern trainees residing in Japan experienced material deprivation in multiple ways. In addition, 23.0% of Vietnamese technical intern trainees in Japan experienced suicidal ideation, indicating a high prevalence of serious psychological problems. Factors contributing to suicidal ideation included age, experience of deprivation in relation to food expenses, and deprivation in relation to cellphone bills. Inability to pay cellphone bills may have increased isolation among Vietnamese trainees in Japan during the COVID-19 pandemic. As the COVID-19 pandemic continues, a system for early detection and support of Vietnamese trainees with suicidal ideation is required. Additionally, it is important to provide financial support for Vietnamese technical intern trainees and prevent their isolation.

## Data Availability

The datasets analyzed in the current study are available from the corresponding author on reasonable request.

## List of abbreviations

CI: confidence interval
COVID-19: coronavirus disease 2019
OR: odds ratio
PHQ-9: Patient Health Questionnaire-9

## Declarations

### Ethics approval and consent to participate

This study was approved by the Kobe City College of Nursing Ethics Research Committee (approval number: 20124-05). Prior to participation in the survey, a statement was made at the beginning of the web page stating the purpose of the study, an overview of the study, that participation in the study was voluntary, that participation was anonymous, and a guarantee that study participants’ information would not be used for any purpose other than scientific research. Those who agreed with the above information were able to participate in the survey by clicking on the consent box. All methods used conformed to the principles and ethical standards of the Declaration of Helsinki.

### Consent for publication

Not applicable

### Author Contributions

T. Y. conceived the original idea for the study. T. Y., P. N. Q., and K. K. designed the questionnaire. T. Y., P. N. Q., and K. K. analyzed the data and wrote the first draft of the manuscript. P. N. Q., C. Y., E. N., E. S., S. I., and K. K. contributed to the interpretation of the results. All authors made substantial intellectual contributions to the study and approved the final draft of the manuscript.

## Acknowledgments

We thank all of the participants who took part in this study. We also thank the faculty of Nursing at Kobe City College of Nursing for their assistance during the ethics submission. We thank Benjamin Knight, MSc., from Edanz (https://jp.edanz.com/ac) for editing a draft of this manuscript.

## Tables

**Table 1.**
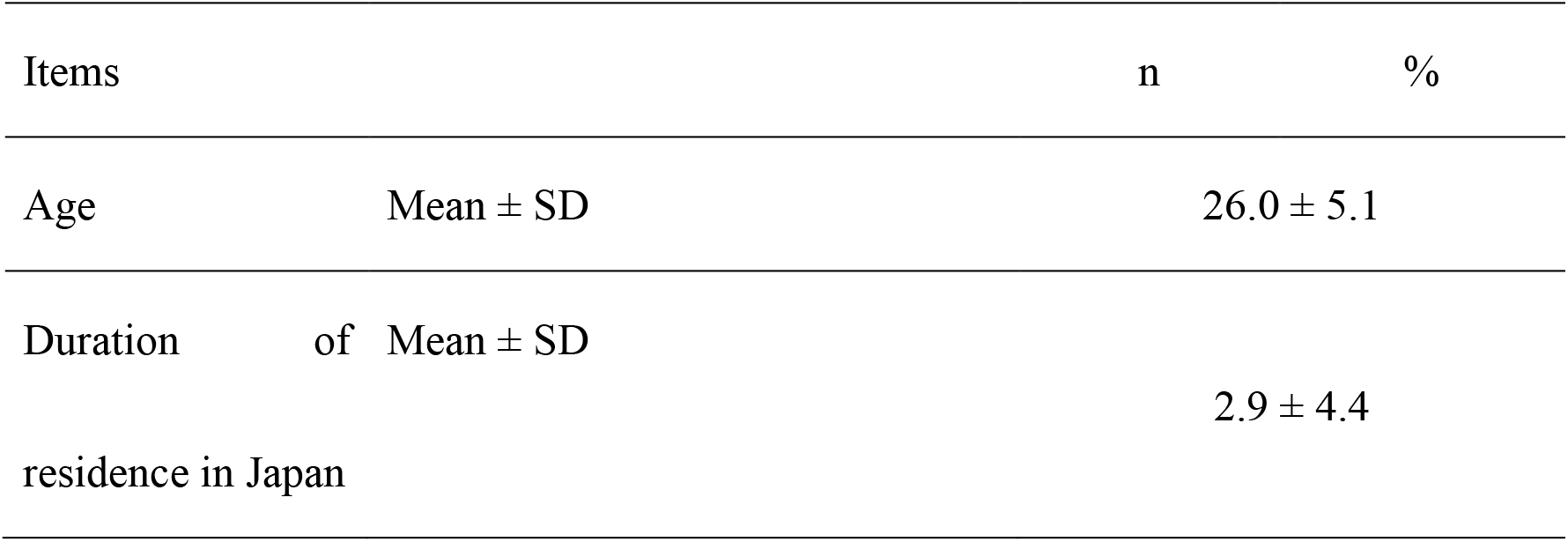
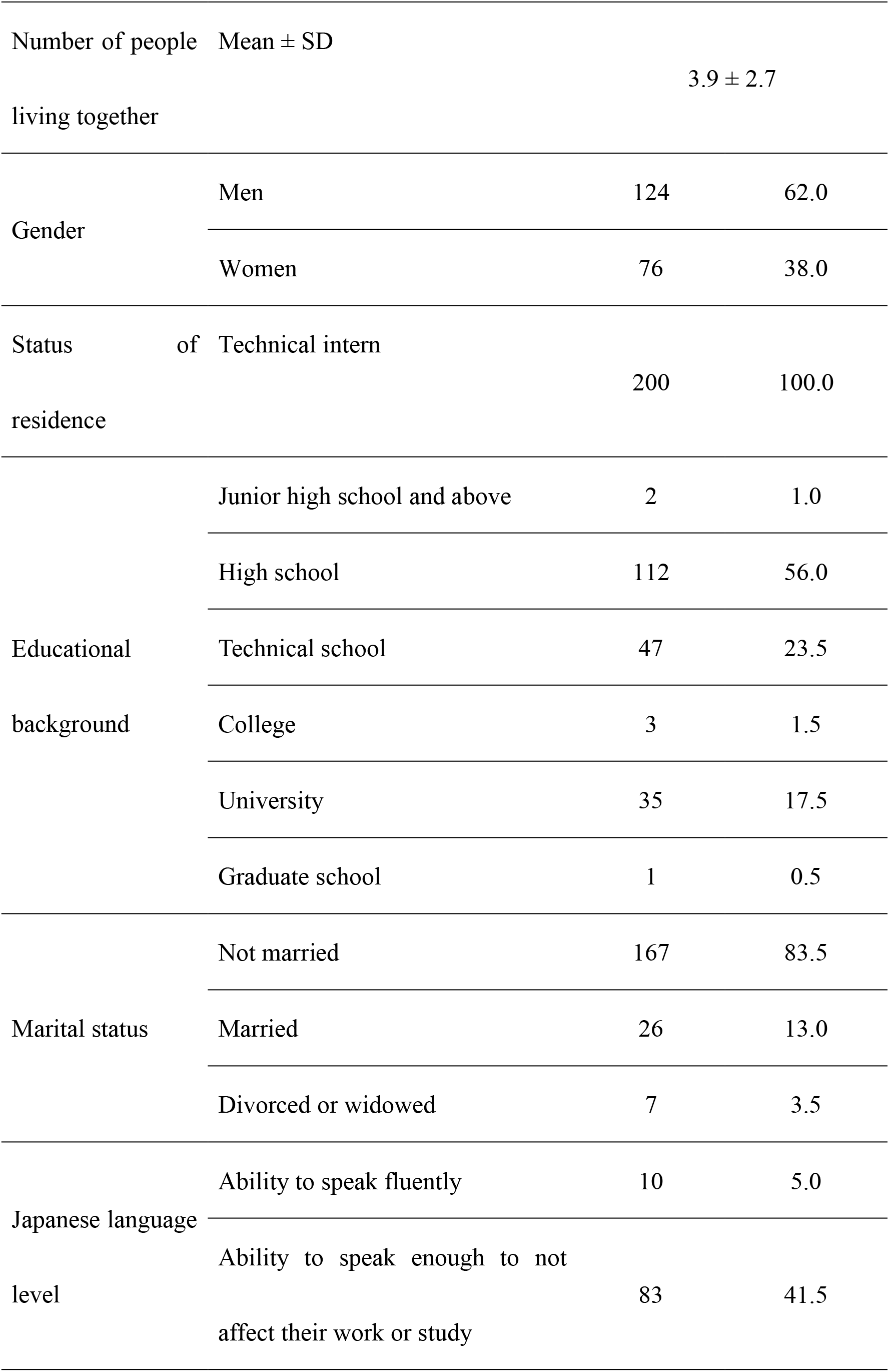
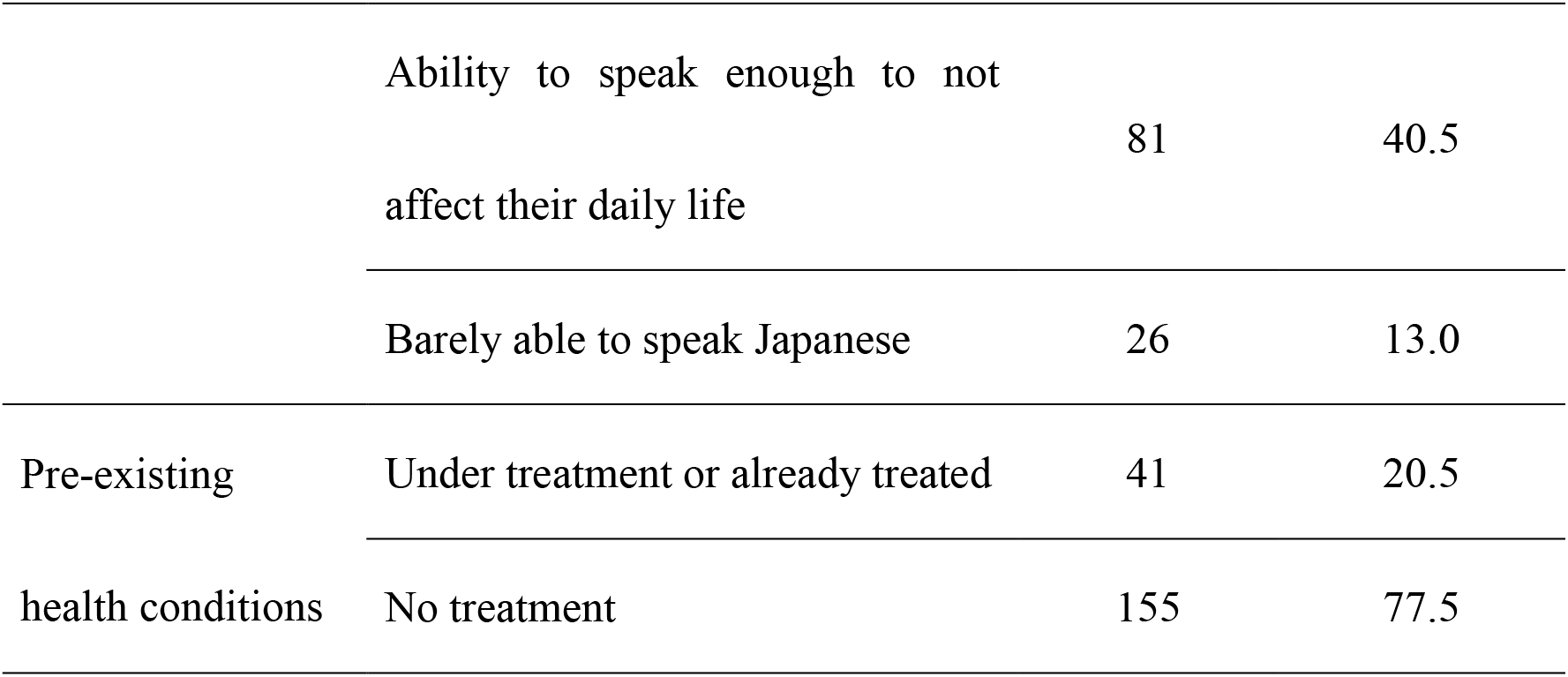
Characteristics of participants (n = 200)

**Table 2.**
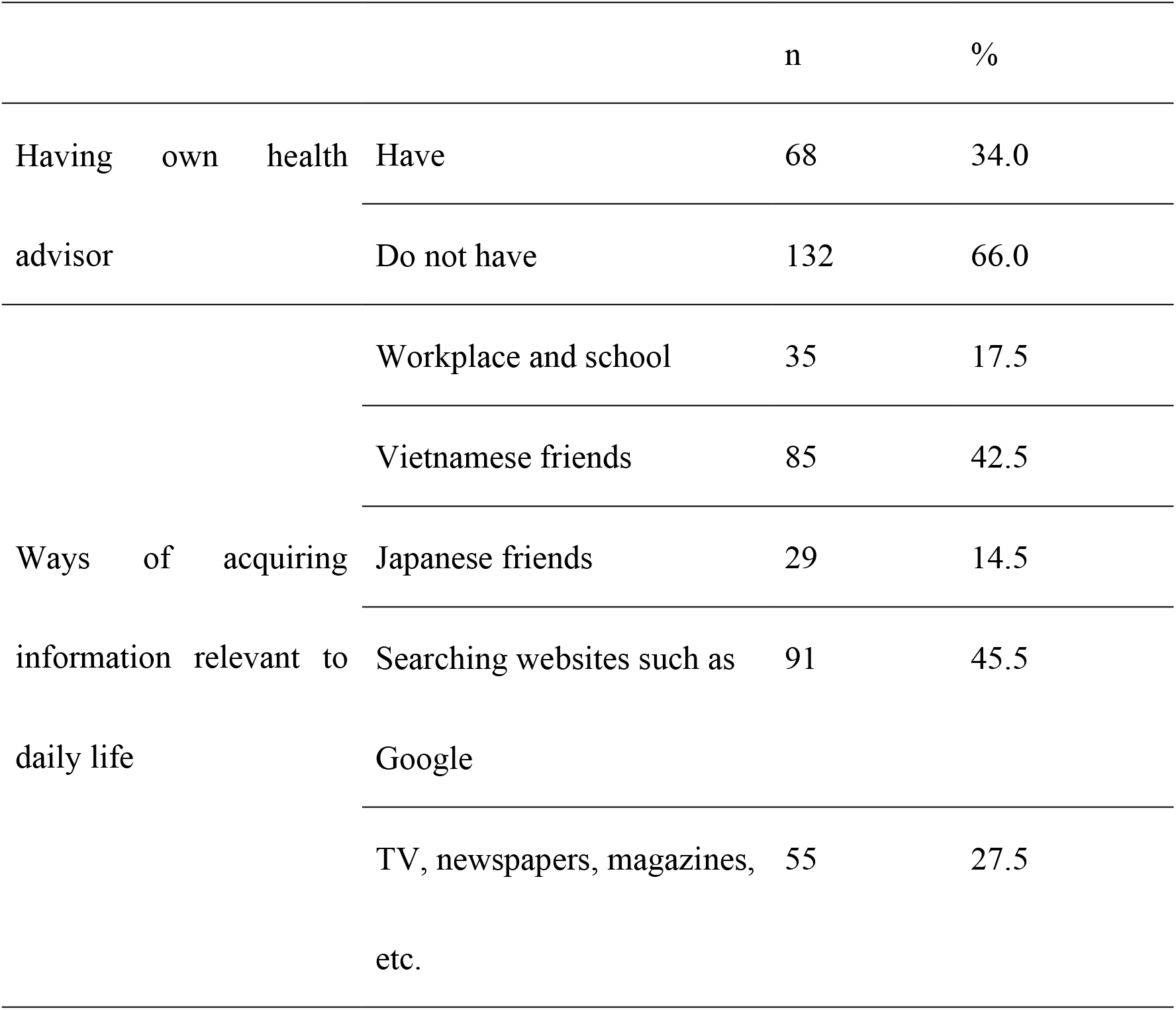
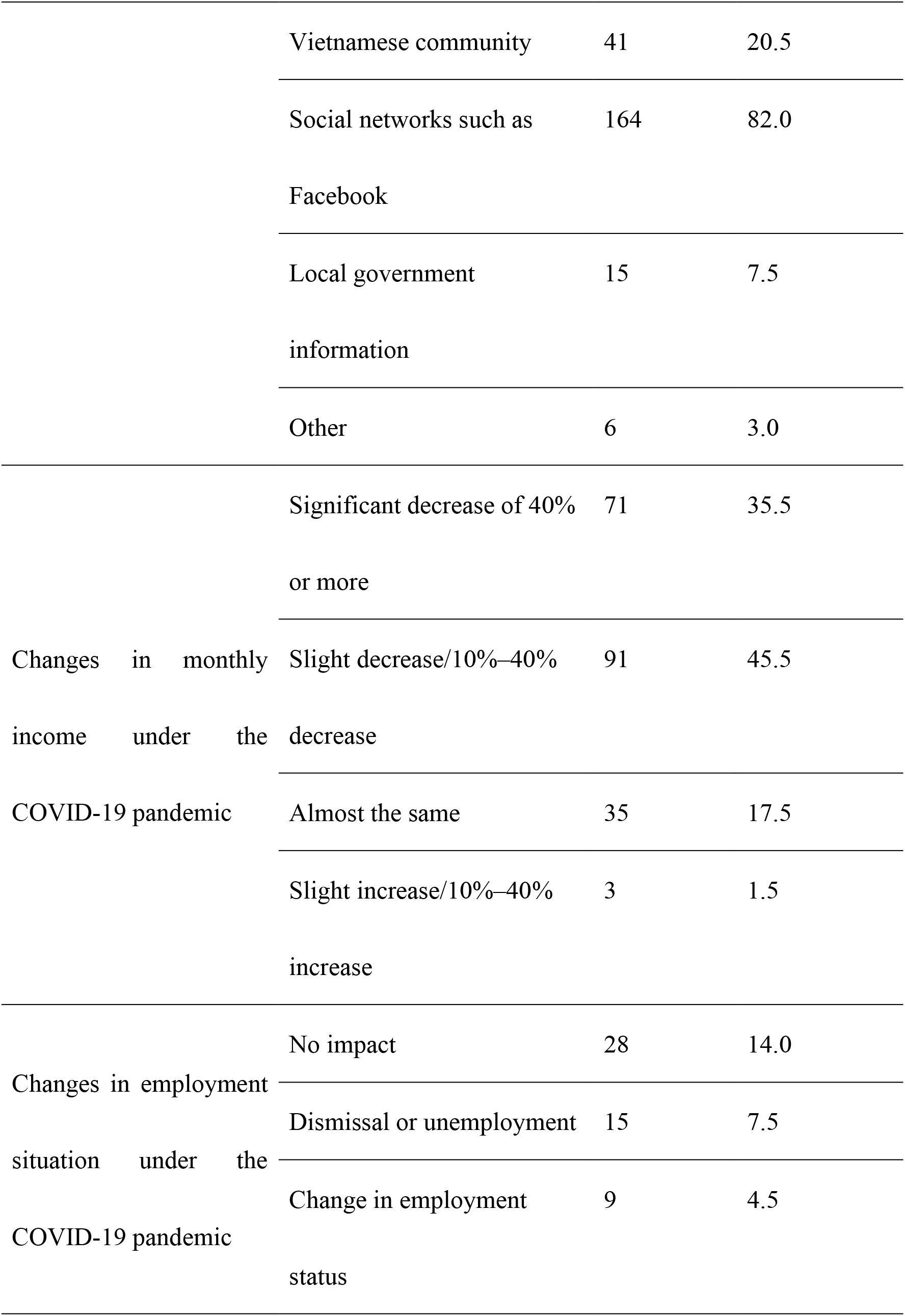
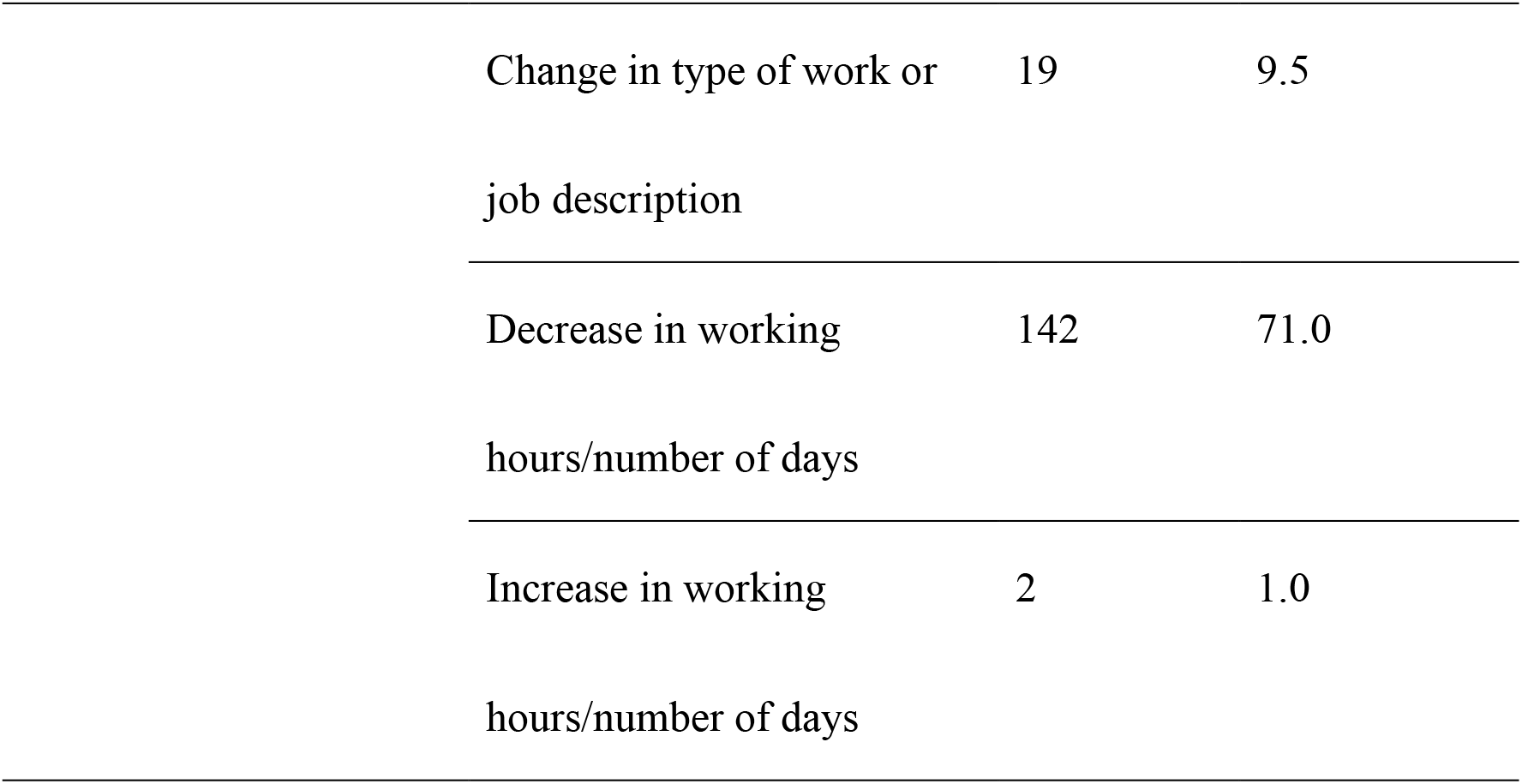
Social and economic situation of Vietnamese technical interns in Japan (n = 200)

**Table 3.**
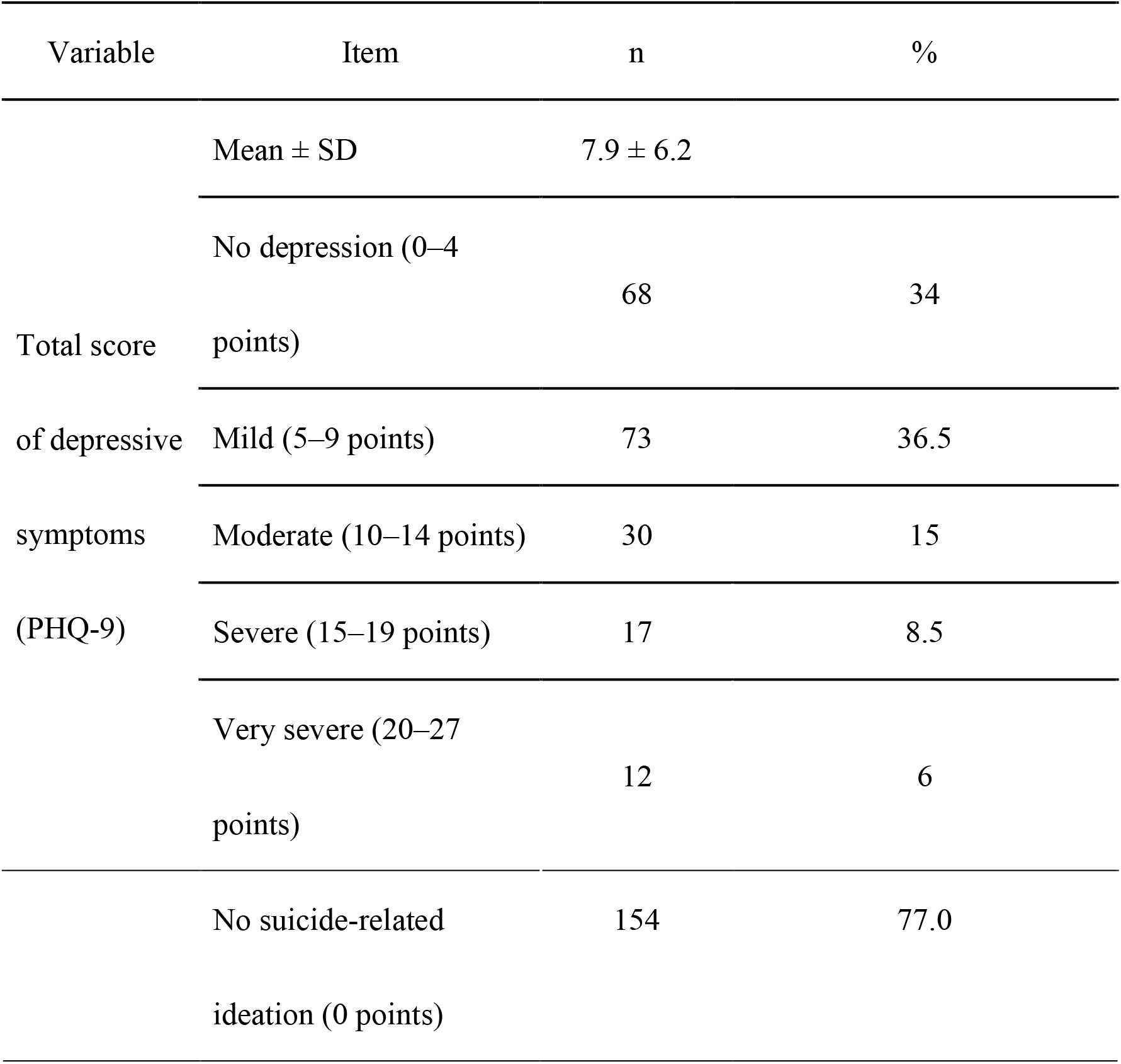
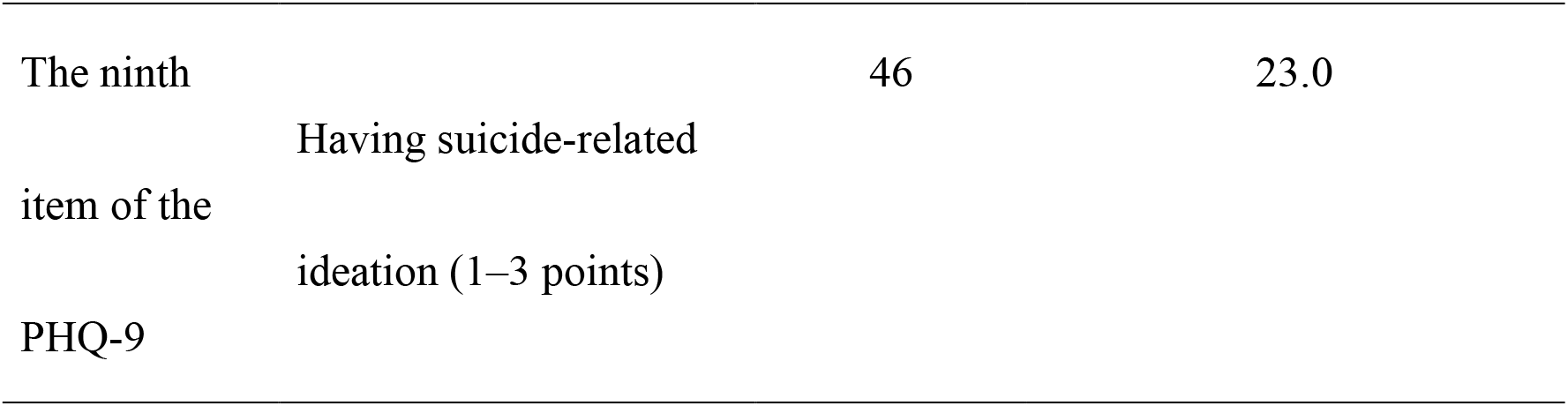
Depression and suicidal ideation among Vietnamese technical interns in Japan

**Table 4.**
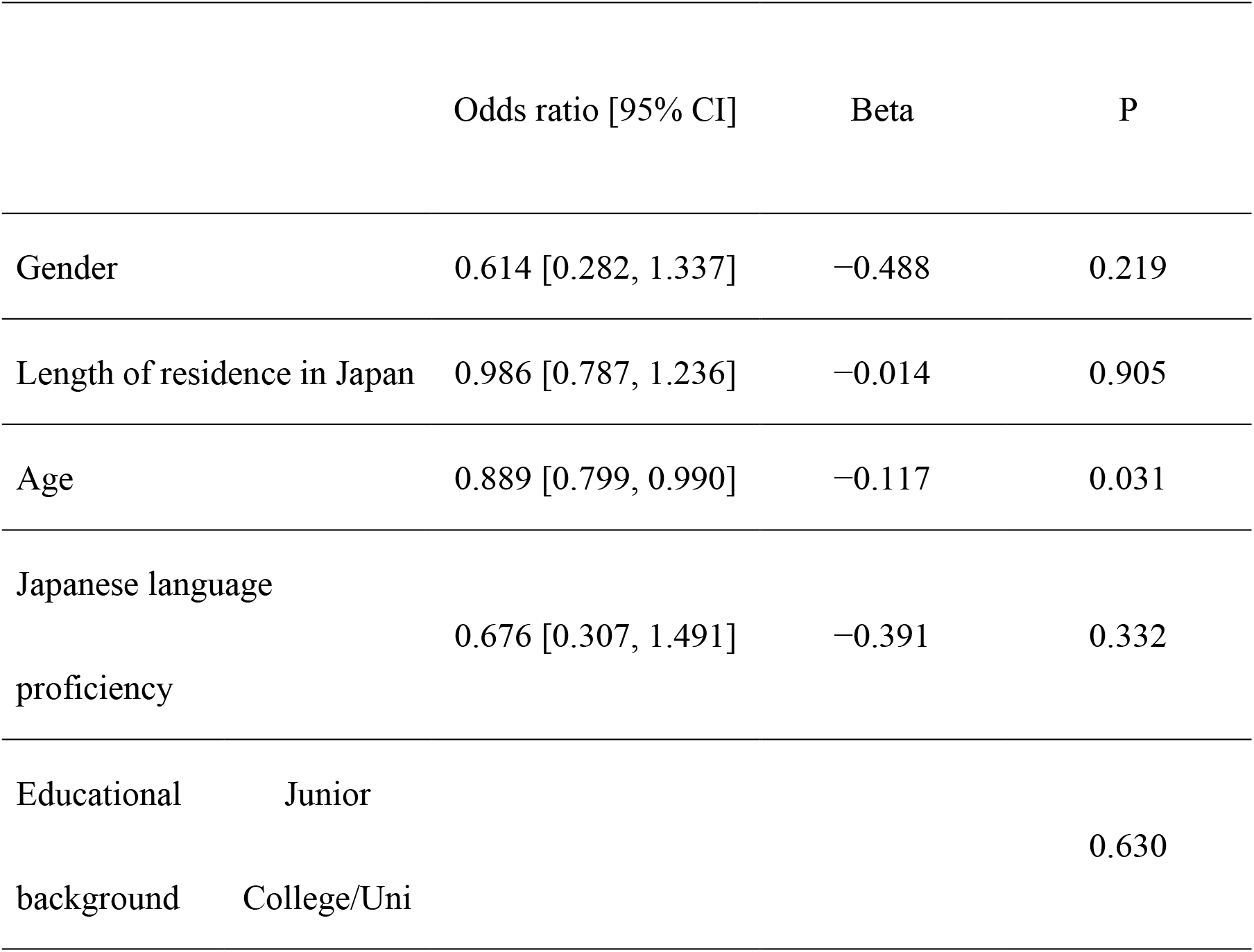
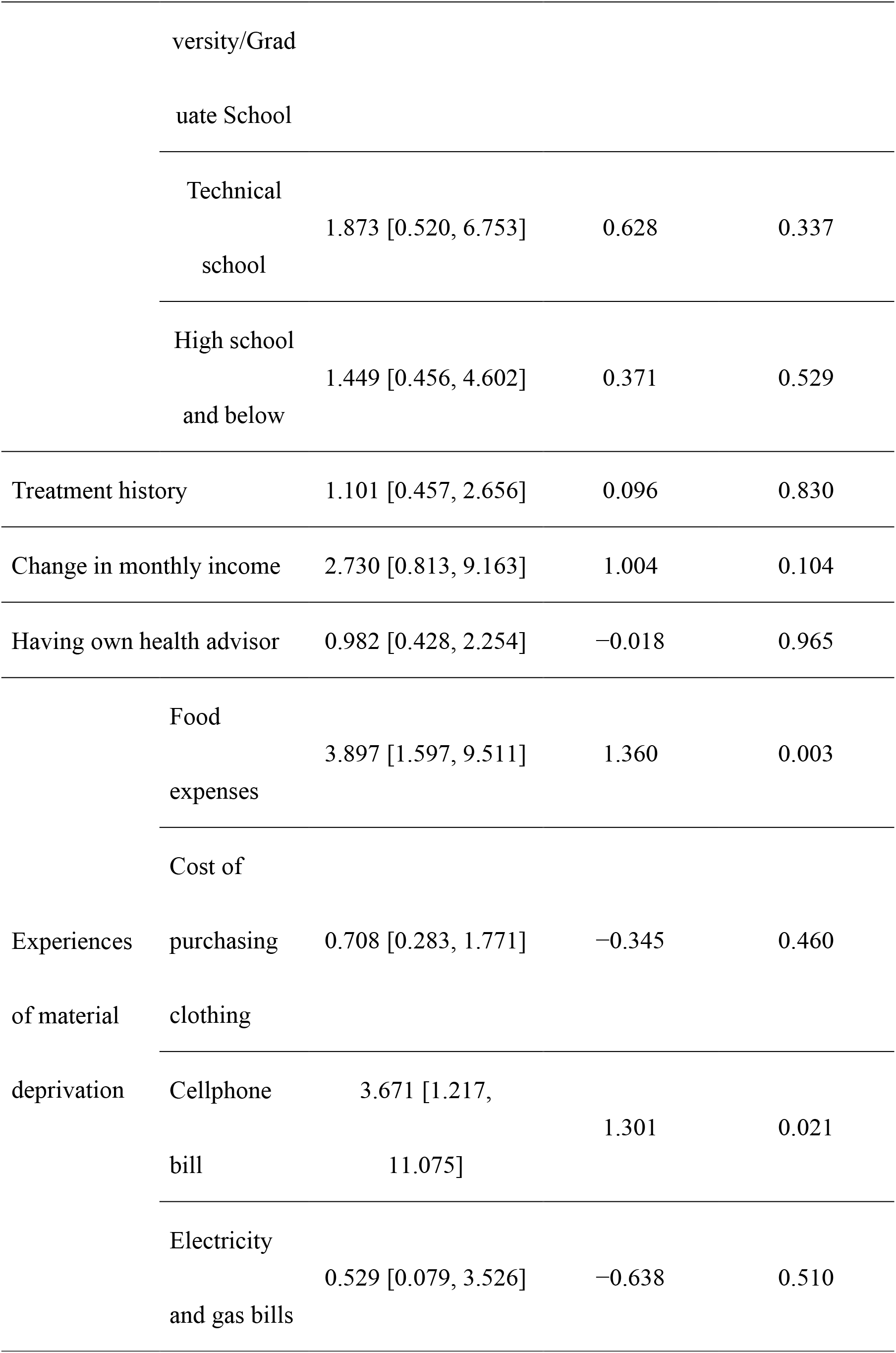
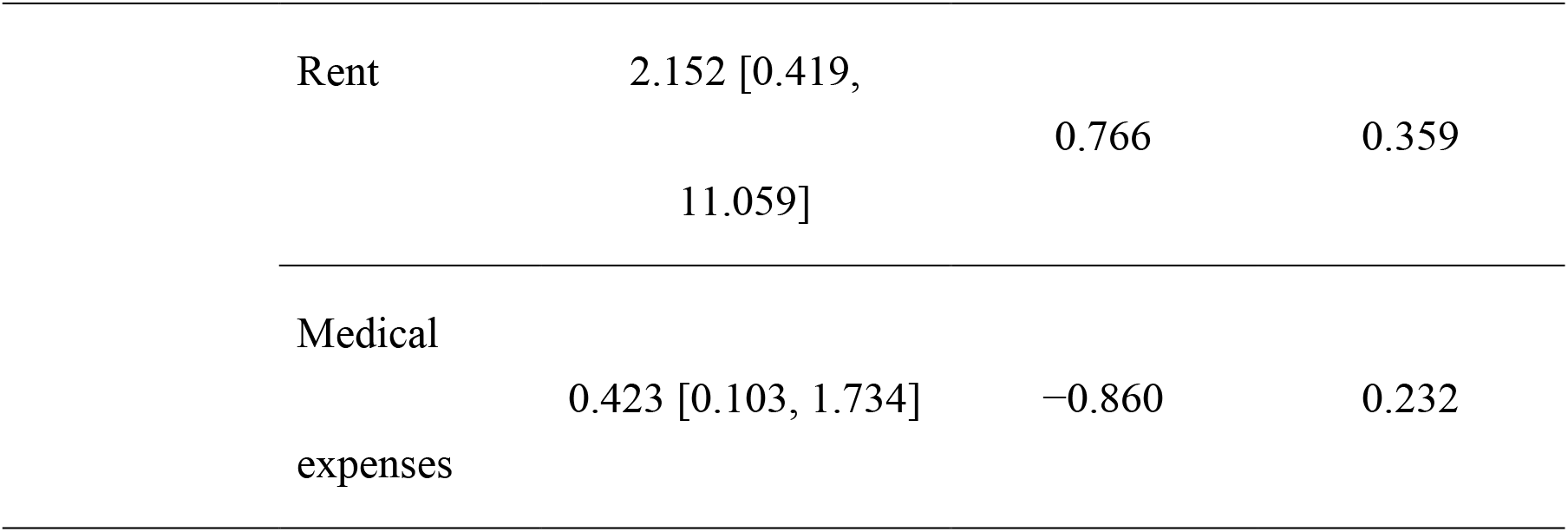
Factors related to suicidal ideation of Vietnamese technical interns in Japan

## References

1. World Health Organization. COVID-19 pandemic triggers 25% increase in prevalence of anxiety and depression worldwide. 2022. https://www.who.int/news/item/02-03-2022-covid-19-pandemic-triggers-25-increase-in-prevalence-of-anxiety-and-depression-worldwide. Accessed 1 Dec 2022.

2. World Health Organization. ApartTogether survey: preliminary overview of refugees and migrants self-reported impact of COVID-19. 2020. https://apps.who.int/iris/bitstream/handle/10665/337931/9789240017924-eng.pdf?sequence=1&isAllowed=y. Accessed 1 Dec 2022.

3. Immigration Services Agency of Japan. Number of foreign residents as of June 30, 2021 (Zairyugaikokujinsu-nisuite). https://www.moj.go.jp/isa/publications/press/13_00017.html (in Japanese). Accessed 1 Dec 2022.

4. Immigration Services Agency of Japan. Recent situations concerning immigration and residency management (Syutunyukokuzanryukanri-womeguru-kinnennojokyou). https://www.moj.go.jp/isa/content/001335866.pdf (in Japanese). Accessed 1 Dec 2022.

5. Immigration Services Agency of Japan. To prevent the disappearance of foreign technical intern trainees (Gaikouzinginouzissyuseino-sissouwo-hasseisasenaitameni). https://www.moj.go.jp/isa/content/001350542.pdf (in Japanese). Accessed 1 Dec 2022.

6. Immigration Services Agency of Japan. Human rights violations against technical intern trainees (Ginouzissyuseinitaisuru-zinkensingai-kouinituite). 2022. https://www.moj.go.jp/isa/content/001363938.pdf (in Japanese). Accessed 1 Dec 2022.

7. Immigration Services Agency of Japan. To supervisory organizations and practical training providers (Kanridantai-zissyuuzissisyanominasamahe). https://www.moj.go.jp/isa/content/001349019.pdf (in Japanese). Accessed 1 Dec 2022.

8. Shinohara A, Kawasaki R, Kuwano N, Ohnishi M. Interview survey of physical and mental changes and coping strategies among 13 Vietnamese female technical interns living in Japan. Health Care for Women Int. 2021; doi:10.1080/07399332.2021.1963966

9. Haisken-DeNew J, Sinning M. Social deprivation of immigrants in Germany. Rev Income Wealth. 2010; doi:10.1111/j.1475-4991.2010.00417.x.

10. Cheung KCK, Chou KL. Poverty, deprivation, and depressive symptoms among older adults in Hong Kong. Aging Ment Health. 2019; doi:10.1080/13607863.2017.1394438.

11. Chung RY, Chung GK, Gordon D, Wong SY, Chan D, Lau MK, et al. Deprivation is associated with worse physical and mental health beyond income poverty: a population-based household survey among Chinese adults. Qual Life Res. 2018; doi:10.1007/s11136-018-1863-y.

12. Tran BQ. Vietnamese technical trainees in Japan voice concerns amidst COVID-19. The Asia-Pacific Journal. 2020;18:5478.

13. Hiroshima Associated Repository Portal. 2021. http://harp.lib.hiroshima-u.ac.jp/h-bunkyo/metadata/12623. Accessed 1 Dec 2022.

14. Bélanger D, Ueno K, Hong KT, Ochiai E. From foreign trainees to unauthorized workers: Vietnamese migrant workers in Japan. Asian Pac Migr J. 2011; doi:10.1177/011719681102000102

15. Sa NTH. The technical intern training program in Japan and Vietnamese trainees. The Bukkyo University Graduate School Review. 2013. https://archives.bukkyo-u.ac.jp/rp-contents/DS/0041/DS00410L019.pdf (in Japanese). Accessed 1 Dec 2022.

16. Cabinet Office, Government of Japan. Chapter 6: Material Deprivation Indicators (bussitutekihakudatusihyou). https://www8.cao.go.jp/kodomonohinkon/chousa/h28_kaihatsu/6_01.html (in Japanese). Accessed 1 Dec 2022.

17. National Institute of Population and Social Security Research. Toward the construction of a deprivation indicator in Japan: an examination of a complementary indicator to the relative poverty rate (nihonniokeruhakudatusihyounokoutikunimukete:soutaitekihinkonrituwohokansurus ihyounokentou). https://www.ipss.go.jp/syoushika/bunken/data/pdf/19919002.pdf (in Japanese). Accessed 1 Dec 2022.

18. Spitzer RL, Williams JBW, Kroenke K. Test review: Patient Health Questionnaire–9 (PHQ-9). Rehabilitation Counseling Bulletin. 2014; doi:10.1177/0034355213515305

19. Pham T, Bui L, Nguyen A, Nguyen B, Tran P, Vu P, Dang L. The prevalence of depression and associated risk factors among medical students: an untold story in Vietnam. PLoS One. 2019; doi:10.1371/journal.pone.0221432.

20. Nguyen TQ, Bandeen-Roche K, Bass JK, German D, Nguyen NTT, Knowlton AR. A tool for sexual minority mental health research: The Patient Health Questionnaire (PHQ-9) as a depressive symptom severity measure for sexual minority women in Viet Nam. J Gay Lesbian Mental Health. 2016; doi:10.1080/19359705.2015.1080204

21. Loerbroks A, Cho SI, Dollard MF, Zou J, Fischer JE, Jiang Y, et al. Associations between work stress and suicidal ideation: individual-participant data from six cross-sectional studies. J Psychosom Res. 2016; doi: https://doi.org/10.1016/j.jpsychores.2016.09.008

22. Chow WS, Schmidtke J, Loerbroks A, Muth T, Angerer P. The relationship between personality traits with depressive symptoms and suicidal ideation among medical students: a cross-sectional study at one medical school in Germany. Int J Env Res Pub Health. 2018; doi:10.3390/ijerph15071462

23. Nomura K, Minamizono S, Maeda E, Kim R, Iwata T, Hirayama J, et al. Cross-sectional survey of depressive symptoms and suicide-related ideation at a Japanese national university during the COVID-19 stay-home order. Environ Health Prev Med. 2021; doi:10.1186/s12199-021-00953-1

24. Mitsui N, Asakura S, Takanobu K, Watanabe S, Toyoshima K, Kako Y, et al. Prediction of major depressive episodes and suicide-related ideation over a 3-year interval among Japanese undergraduates. PLOS ONE. 2018; doi:10.1371/journal.pone.0201047

25. Hovey JD. Acculturative stress, depression, and suicidal ideation among Central American immigrants. Suicide Life Threat Behav. 2000; doi:10.1111/j.1943-278X.2000.tb01071.x

26. Hovey JD. Acculturative stress, depression, and suicidal ideation in Mexican immigrants. Cultur Divers Ethnic Minor Psychol. 2000; doi:10.1037/1099-9809.6.2.134.

27. Beutel ME, Jünger C, Klein EM, Wild P, Lackner KJ, Blettner M, et al. Depression, anxiety and suicidal ideation among 1st and 2nd generation migrants: results from the Gutenberg health study. BMC Psychiatry. 2016; doi:10.1186/s12888-016-0995-2.

28. Fortuna LR, Álvarez K, Ramos Ortiz Z, Wang Y, Mozo Alegría X, Cook BL, et al. Mental health, migration stressors and suicidal ideation among Latino immigrants in Spain and the United States. Eur Psychiatry. 2016; doi:10.1016/j.eurpsy.2016.03.001.

29. Immigration Services Agency of Japan. Support for maintaining employment for technical intern trainees who have difficulty continuing practical training due to the impact of new coronavirus infection (Singatakoronauirusukannsennsyounoeikyouniyori-zissyuugakeizokukonnantonatta-ginouzissyuuseitounitaisuru-koyouizisien). https://www.moj.go.jp/isa/nyuukokukanri14_00008.html (in Japanese). Accessed 1 Dec 2022.

30. Catholic Commission of Japan for Migrants, Refugees and People on the Move (Nihonkatorikku-nannmin-ijuuidousha-iinkai). https://www.jcarm.com/ (in Japanese). Accessed 22 Oct 2022.

31. Catholic Commission of Japan for Migrants, Refugees and People on the Move. Technical intern hotline report - increasingly complex issues and the possibility of “resistance” (Ginouzissyuusei-hottorainhoukoku). https://www.jcarm.com/wordpress/wp-content/uploads/2020/10/20200812hotline_report.pdf (in Japanese). Accessed 22 Oct 2022.

32. Yasukawa K, Sawada T, Hashimoto H, Jimba M. Health-care disparities for foreign residents in Japan. The Lancet. 2019; doi:10.1016/S0140-6736(19)30215-6

33. Thi HT, Kitajima T, Takeshi S, Hiroko M. Mental health and associated factors for Vietnamese migrants in Japan during the COVID-19 pandemic: a comparative analysis on resident status. 2022; doi:10.21203/rs.3.rs-1552849/v1

34. Ishimaru T, Kuraoka H, Shimizu S, Hara K. Current status and challenges of supervisory organizations’ support for the health and safety of technical intern trainees: focusing on collaboration with occupational health professionals. J Occup Health. 2021; doi:10.1539/sangyoeisei.2021-050-B

35. Pak TY, Choung Y. Relative deprivation and suicide risk in South Korea. Soc Sci Med. 2020; doi:10.1016/j.socscimed.2020.112815

36. Huang Q, Lin S, Li Y, Huang S, Liao Z, Chen X, et al. Suicidal ideation is associated with excessive smartphone use among Chinese college students. Front Public Health. 2022; doi:10.3389/fpubh.2021.809463

37. Li G, Conti AA, Qiu C, Tang W. Adolescent mobile phone addiction during the COVID-19 pandemic predicts subsequent suicide risk: a two-wave longitudinal study. BMC Public Health. 2022; doi:10.1186/s12889-022-13931-1

38. Nguyen TXT, Lal S, Abdul-Salam S, Yuktadatta P, McKinnon L, Khan MSR, et al. Has smartphone use influenced loneliness during the COVID-19 pandemic in Japan? Int J Env Res Pub Health. 2022; doi:10.3390/ijerph191710540

39. Cauberghe V, Van Wesenbeeck I, De Jans S, Hudders L, Ponnet K. How adolescents use social media to cope with feelings of loneliness and anxiety during COVID-19 lockdown. Cyberpsych Behav Soc Net. 2021; doi:10.1089/cyber.2020.0478

40. Li SH, Beames JR, Newby JM, Maston K, Christensen H, Werner-Seidler A. The impact of COVID-19 on the lives and mental health of Australian adolescents. Eur Child Adolesc Psychiatry. 2022; doi:10.1007/s00787-021-01790-x.

